# CD14 (C-159T) polymorphism is associated with increased susceptibility to SLE, and plasma levels of soluble CD14 is a novel biomarker of disease activity: a hospital-based case-control study

**DOI:** 10.1101/2020.06.19.20136119

**Authors:** Aditya K Panda, Rina Tripathy, Bidyut K Das

## Abstract

**Background:** Cluster of differentiation 14 (CD14) plays a crucial role in the innate immune response of the host in protection against various pathogens. The importance of soluble CD14 in autoimmune disorders has been described in different populations. However, the role of sCD14 in systemic lupus erythematosus (SLE) is poorly understood. Further, the association of functional variants at the promoter region of the CD14 gene (−159 C>T) with susceptibility to SLE or disease severity needs to be defined.

**Methods:** Two hundred female SLE patients diagnosed on SLICC classification criteria and age, sex, matched healthy controls were enrolled in the present study. PCR-RFLP method was used to genotype CD14 (C-159 T) polymorphism. Plasma levels of IFN-α, TNF-α, and sCD14 were quantified by ELISA.

**Results:** Prevalence of mutant genotypes (CT and TT) and minor allele of CD14 (C-159T) polymorphism was significantly higher in SLE cases compared to healthy controls (CT:P<0.0001; OR=3.26, TT:P<0.0001; OR=3.39; T:P=0.0009, OR=1.62). Further, lupus nephritis patients had a higher prevalence of homozygous mutants (TT) and mutant allele (T)(TT: P=0.0002, OR=8.07; T: P=0.001, OR=1.32). SLE patients displayed significantly increased plasma sCD14, TNF-α, and IFN-α levels in comparison to healthy controls. These cytokines were significantly elevated in patients of lupus nephritis compared to those without kidney involvement. Interestingly, sCD14 levels correlated positively with SLEDAI-2K scores and 24 hours proteinuria.

**Conclusion:** CD14 (C-159T) polymorphism is associated with an increased predisposition to the development of SLE and lupus nephritis: sCD14 is a promising novel biomarker for assessing disease activity and lupus nephritis.

## INTRODUCTION

The systemic lupus erythematosus (SLE) is a complex, autoimmune disease characterized by chronic inflammation in various tissues of the body(1). Although the exact mechanism of induction of autoimmune diseases is unknown, dysregulation of the adaptive immune system has been attributed to autoantibody production(1). Both innate and adaptive immune systems closely interact with one another, with adaptive immunity being controlled by the components of innate immunity (2). The innate immune system can be triggered by various infectious agents which possibly play a significant role in the pathogenesis of SLE(3).

Toll-like receptors (TLRs), present on the cell surface of innate immune cells, recognize microbial motifs called pathogen-associated molecular patterns (PAMPs) and activate the downstream signal transduction pathways(4). There are several lines of evidence to suggest that TLR4 by itself cannot act as a receptor for lipopolysaccharide (LPS) for the induction of downstream signaling. It requires other molecules viz. LPS-binding protein (LBP), myeloid differentiation protein 2 (MD-2), and CD14 for effective signaling (5). The critical role of CD14 in LPS signaling has been well documented - CD14 knockout mice are resistant to LPS challenge (6). The addition of sCD14 enhanced TLR4 mediated NFκ-B activation in the HEK-293 cell line (7).

Since CD14 plays a crucial role in the innate immune system and infections have been an important triggering factor for the development of SLE, a small study reported by Nockher et al in 1994 showed elevated sCD14 levels in patients of SLE; active SLE patients displayed significantly higher levels compared to patients with inactive disease. Furthermore, sCD14 levels correlated positively with disease activity scores (8).

Differential levels of sCD14 have been reported in healthy individuals of different populations. Variations in plasma sCD14 levels have been attributed to the genetic makeup of the subjects and associated with various functional gene polymorphisms. A polymorphism at the promoter region (−159) leading to a C-to-T change at that position was associated with increased CD14 mRNA expression (9). Homozygous mutants (TT) were associated with higher levels of soluble and membrane-bound CD14 (10, 11). Since CD14 (C-159T) polymorphism up-regulates levels of sCD14, we hypothesized that the variant might be associated with an increased predisposition to SLE as well as to its severity. We addressed this hypothesis by enrolling SLE patients admitted in a tertiary care center and healthy controls from Odisha, an Eastern state of India; CD14 (−159) polymorphism was genotyped, plasma levels of sCD14 inflammatory cytokines were quantified and analyzed for association with SLE and its disease severity (SLEDAI-2K).

## Materials and Methods

### Patients and controls

SLE is a chronic inflammatory disorder and mostly affects females(12). Female SLE patients attending rheumatology clinic and/or admitted to SCB Medical College, Cuttack, Odisha, India, were enrolled in the present study. The diagnosis of SLE was based on the SLICC 2012 classification criteria(13). After a detailed clinical examination and laboratory investigations, patients were categorized under different clinical phenotypes. The clinical profile of 200 SLE patients has been summarized in Table-1. The disease severity was recorded by the SLEDAI-2K scores. Two hundred, age-matched females were included as healthy controls (HC). None of the controls reported a history of autoimmune disorder. About 5 ml of blood in EDTA was collected; plasma was separated and quantified for sCD14, TNFα, and IFNα levels by enzyme-linked immunosorbent assay (ELISA). The remaining blood samples were preserved with an equal amount of 8M urea at -20^°^C until use for isolation of DNA. The study was approved by the Institutional Ethics Committee of SCB Medical College Cuttack, Odisha, India. Informed written consent was obtained from each patient.

**Table 1.** Clinical characteristics of SLE patients and healthy controls.

| Clinical profiles | SLE (*n*=200) | Healthy control (*n*=200) |
| --- | --- | --- |
| Sex (male/ female) | 0/200 | 0/200 |
| Age (mean ±SD) | 28.23±8.48 | 29.24 ±6.48 |
| Duration of disease (mean ±SD) | 1.67 ±2.32 | - |
| Clinical Profile: |  |  |
| Photosensitivity | 48 (24) | - |
| Malar rash | 71 (35.5) | - |
| Discoid rash | 34 (17) | - |
| Oral ulcer | 80 (40) | - |
| Arthritis | 88 (44) | - |
| NPSLE | 37 (18.5) | - |
| Myocarditis | 18 (9) | - |
| AIHA | 32 (16) | - |
| Serositis | 36 (18) | - |
| Nephritis | 104 (52) | - |
| Pneumonitis | 15 (7.5) | - |

Note. Data are no. (%) of participants unless otherwise specified. NPSLE, Neuropsychiatric systemic lupus erythematosus; AIHA, autoimmune hemolytic anemia.

### Genotyping of CD14 (C-159T) polymorphism

Genomic DNA was extracted using the QIAamp DNA Blood Mini Kit (Qiagen, Hilden, Germany) according to the manufacturer’s instructions. PCR followed by restriction fragment length polymorphism (PCR-RFLP) technique was employed to type the polymorphism at the promoter region (C-159T) of the CD14 gene (14). In brief, primer set (forward primer-5’-GCC TCT GAC AGT TTA GTA ATC –3’ and reverse primer-5’-GTG CCA ACA GAT GAG GTT CAC –3’) was used for amplification of CD14 promoter region flanking C-159T polymorphic region which produced an amplicon of size 497bp. The amplified product was digested by restriction enzyme *HaeIII* and based on the differential product size individuals genotypes for CD14 (C-159T), polymorphism was determined (CC: 198+156+143bp; TT: 299+198bp and CT: 299+198+156+143bp). Subsequently, 50 samples were randomly selected and genotyped through direct sequencing. The result of the sequencing and PCR-RFLP method was found to be in concordance.

### Determination of plasma sCD14, IFN-α, and TNF-α

The plasma samples of 46 healthy controls and 78 SLE patients were available; sCD14, IFN-α and TNF-α levels were quantified by ELISA according to the manufacturer’s instructions (sCD14 kit from R&D Systems, Wiesbaden, Germany, IFN-α and TNF-α from Bender Med Systems, Vienna, Austria).

### In silico analysis

Two online tools were used to investigate the functional relevance of CD14 promoter polymorphism (C-159T)) i) SNPinfo (FuncPred), and ii) RegulomeDB. As described earlier (15), the reference id of SNPs was entered into the online program of SNPinfo (FuncPred) (https://snpinfo.niehs.nih.gov/snpinfo/snpfunc.html) with the default setting. The output information was noted down for the possible functional effect of the genetic mutation.

To further strengthen the result of SNPinfo, another programRegulomeDB (http://regulomedb.org/)(16) was employed to investigate the functional relevance of CD14 promoter polymorphism. The result of the RegulomeDB categorizes into six groups (score-1: likely to affect binding and linked to the expression of a gene target; score-2: likely to affect binding; score-3: less likely to affect binding; score 4,5 and 6: minimal binding evidence). With no annotation data available, RegulomeDB allocates a score of 7. In the present study, dbSNP ID was used for the analysis.

### Statistical analysis

Genotype and allele frequency was calculated by direct counting. SNPalyze software (Dynacom, Japan) was employed to calculate the Hardy-Weinberg equilibrium. Fisher’s test was used for comparison of genotype, allele frequencies between SLE patients and healthy controls Wild type (CC) and major allele (C) were selected as reference (OR=1) and the other ORs and 95% confidence interval were calculated relative to that reference (Fisher’s exact test, 2×2 contingency tables). A P value of less than 0.01 was taken as significant. Distribution of plasma sCD14, IFN-α, and TNF-α in different CD14 (C-159T) genotypes and clinical categories were assessed by D’Agostino & Pearson omnibus normality test. Based on the results of the normality test, the association of genotype with plasma sCD14, TNF-α, and IFN-αwere analyzed by ANOVA or Kruskal-Wallis test followed by Tukey’s post-test. The Mann-Whitney analysis was employed to compare the mean plasma levels of sCD14, TNF-α, and IFN-α in controls and SLE patients, or different clinical categories of SLE patients. All correlation analysis was performed by Spearman rank correlation test, and *P* value less than 0.05 were considered as significant. Graphpad prism 7.05 software was used for these statistical analyses.

## Results

### Baseline characteristics

A total of 200 female SLE patients and the same number of healthy controls were enrolled in the current study. Baseline data are shown in Table-1. The age profile of patients and healthy controls were comparable. All patients were naïve, and the mean disease duration was 1.67 years. Similar to our earlier reports(17-19), renal involvement was observed in a majority of patients (52%) followed by arthritis (44%) and oral ulcer in (40%) of patients.

### CD14 (C-159T) polymorphism is associated with SLE and lupus nephritis

An association of CD14 (C-159T) polymorphism with SLE and its clinical manifestation was assessed. As shown in Table-2, the prevalence of homozygous mutant (TT) and heterozygous (CT) was significantly higher in SLE patients compared to healthy controls (TT: *P*=0.0001, OR=3.39, 95% CI=1.76 to 6.22; CT: *P*<0.0001, OR=3.26, 95% CI=1.85 to 5.82). Furthermore, the mutant allele (T) was also significantly associated with SLE (*P*=0.0009, OR=1.62, 95% CI=1.22 to 2.13) (Table-2).

**Table 2.** Prevalence of CD14 (C-159T) polymorphism in SLE patients and healthy controls.

| Genotype/Allele | HC (n=200) | SLE (n=200) | P value | OR (95% CI) |
| --- | --- | --- | --- | --- |
| Genotype |  |  |  |  |
| CC | 58 (29) | 22 (11) | 1 | ref |
| CT | 100 (50) | 124 (62) | <0.0001 | 3.26 (1.85 to 5.82) |
| TT | 42 (21) | 54 (27) | 0.0001 | 3.39 (1.76 to 6.22) |
| Allele |  |  |  |  |
| C | 216 (54) | 168 (42) | 1 | Ref |
| T | 184 (46) | 232 (58) | 0.0009 | 1.62 (1.22 to 2.13) |

Note. Data are no. (%) of participants unless otherwise specified. HC: healthy control; SLE: systemic lupus erythematosus; OR: odds ratio; 95% CI: 95% confidence interval. Distribution of genotypes and allele were compared by Fisher exact test.

We analyzed whether CD14 promoter polymorphism was associated with clinical manifestations of SLE. As shown in Table-3, the frequency of homozygous mutant (TT) was significantly higher in patients with lupus nephritis compared to those without renal involvement (*P*=0.0002, OR=8.07, 95% CI=2.44 to 22.53). Distribution of mutant allele also showed a similar association (*P*=0.001, OR=1.32, 95% CI=1.22 to 2.96). However, the presence of CD14 (C-159T) polymorphism was comparable among other clinical categories (data not shown).

**Table 3.** Distribution of CD14 (C-159T) polymorphism in SLE patients with lupus nephritis.

| Genotype/Allele | LN<sup>-</sup> (n=96) | LN<sup>+</sup> (n=104) | *P* value | OR (95% CI) |
| --- | --- | --- | --- | --- |
| Genotype |  |  |  |  |
| CC | 17 (18) | 5 (4) | 1 | ref |
| CT | 63 (66) | 61 (59) | 0.03 | 3.29 (1.23 to 8.49) |
| TT | 16 (16) | 38 (37) | 0.0002 | 8.07 (2.44 to 22.53) |
| Allele |  |  |  |  |
| C | 97 (51) | 71 (34) | 1 | ref |
| T | 95 (49) | 137 (66) | 0.001 | 1.32 (1.22 to 2.96) |

Note. Data are no. (%) of participants unless otherwise specified. LN<sup>+</sup>: patients with lupus nephritis; LN<sup>-</sup>: patients without lupus nephritis manifestation; OR: odds ratio; 95% CI: 95% confidence interval. Distribution of genotypes and allele were compared by Fisher exact test.

### Functional relevance of CD14(C-159T) polymorphism

To determine the functional relevance of CD14 (C-159T) polymorphism, plasma sCD14, IFN-α, and TNF-α levels were assessed in healthy controls and SLE patients (CC=19; CT=73; TT=32). As shown in Figure-1A, homozygous mutant (TT) displayed significantly higher plasma sCD14 levels in comparison to the wild type (CC) (*P*<0.0001) and heterozygous mutant (CT) (*P*=0.001). Besides, homozygous mutant (TT) for CD14 (C-159T) polymorphism also displayed significantly higher plasma IFN-α (*P*<0.0001) (Figure-1B) and TNF-α (*P*<0.0001) (Figure-1C) compared to the wild types (CC).

**Figure-1.**
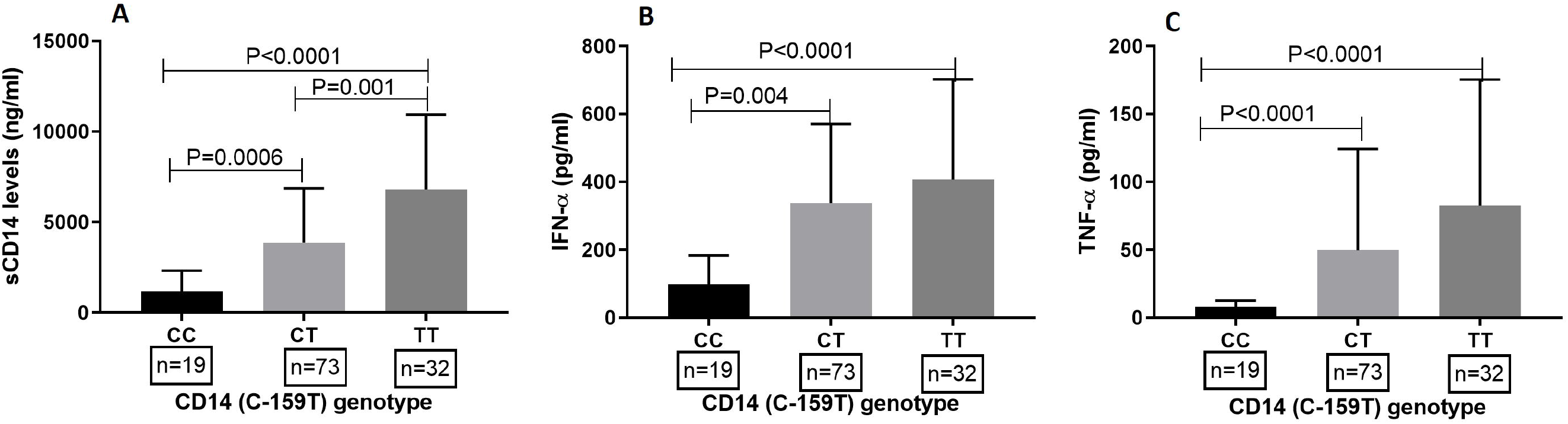
Association of CD14(C-159T) polymorphism with plasma sCD14, IFN-α and TNF-α. Plasma concentrations of sCD14, IFN-α and TNF-α in healthy controls and SLE patients were measured by commercial kit. Based on availability plasma, 78 SLE patients (SLE) and 46 healthy controls (HC) were quantified for sCD14 (A), IFN-α (B) and TNF-α (C) and correlated with CD14 (C-159T) polymorphism. Numbers of samples from each genotype are shown in box. Mean plasma levels in different genotypes were compared by Analysis of Variance (ANOVA) or Kruskal-Wallis test as appropriate followed by Turkey post-test. *P* value less than 0.05 was considered as significant.

### Functional relevance of CD14 (C-159T) polymorphism: in silico

We observed a significant association between plasma levels of sCD14 and CD14 promoter polymorphism (C-159T). SNPinfo (FuncPred) and RegulomeDB online tools were used for validation of the above result in silico, and the output data are shown in Supplementary Table-1. FuncPred program revealed a significant role of CD14 (C-159T) polymorphism on affecting the binding sites of transcription factors (TFBS). However, variation at -159 sites did not affect the miRNA binding location and found to have a regulatory potential (RegPot) of 0.26.

The functional relevance of CD14 polymorphism (C-159T) analysis with RegulomeDB revealed a score of ‘1b’ which indicates a likely binding effect and which is linked to the expression of a gene target. Furthermore, it had annotation for eQTL + TF binding + any motif + DNase footprint + DNase peak, indicating a decisive role in the regulation of the CD14 gene (Supplementary Table-1).

### SLE patients and those with lupus nephritis had higher plasma sCD14, IFN-α and TNF-α

We demonstrated previously that the prevalence of the TT genotype was significantly higher in SLE patients and those with lupus nephritis. And this genotype (TT) was associated with higher plasma sCD14, IFN-α, and TNF-α levels These observations alluded us to investigate the association of plasma levels of sCD14, IFN-α, and TNF-α in SLE to disease manifestations. As shown in Figure-2, plasma levels of sCD14 (A), IFN-α (B) and TNF-α (C) were significantly higher in SLE patients compared to healthy controls (*P*<0.0001). To assess the association of plasma parameters such as sCD14, TNF-α, and IFN-α with lupus nephritis, SLE patients were divided into two group i) patients with lupus nephritis (n=38) and ii) patients without lupus nephritis (n=40). Subjects with lupus nephritis displayed significantly higher plasma sCD14 (D), IFN-α (E) and TNF-α (F) compared to the other group (sCD14: *P*<0.0001, IFN-α: *P*=0.001, TNF-α: *P*=0.005) (Figure-2).

**Figure-2.**
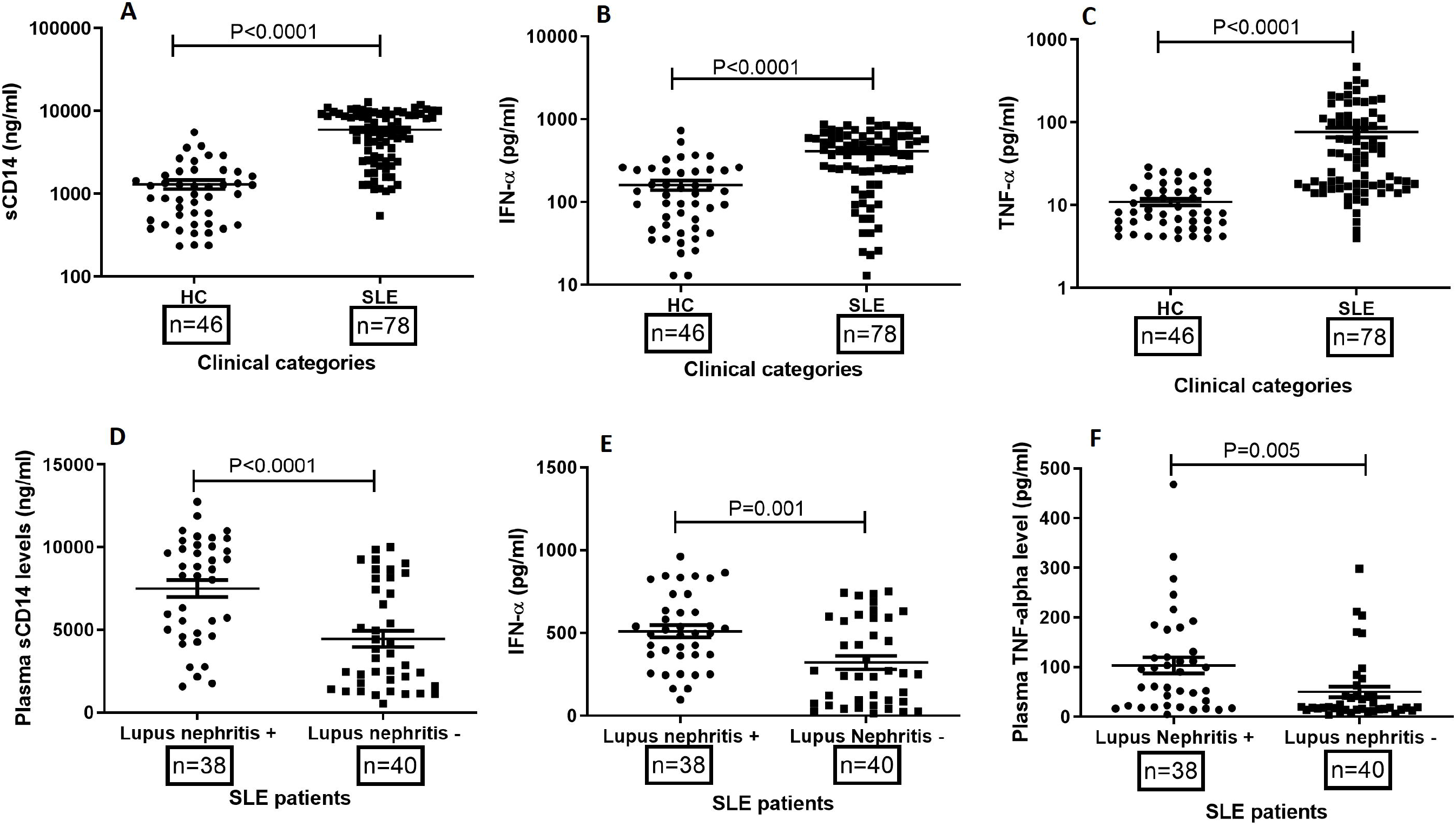
Plasma levels of sCD14, IFN-α and TNF-α in SLE patients. Plasma samples of healthy controls (n=46) and SLE patients (n=78) were quantified for sCD14, IFN-α and TNF-α levels by ELISA according to manufacturer’s instructions. SLE patients displayed significantly higher concentrations of sCD14 (A), IFN-α (B) and TNF-α (C) compared to healthy controls. Based on kidney dysfunction clinical phenotype, SLE patients were further categorized in to lupus nephritis^+^ and lupus nephritis^-^. Lupus patients with nephritis displayed significantly higher sCD14 (D), IFN-α (E) and TNF-α (F) compared to those free of kidney dysfunction. Dots represent individual samples; bars show the mean ± SEM. Mann-Whitney test was used to compare plasma concentrations among different clinical categories. *P* values less than 0.05 was considered as significant.

### 24-hour proteinuria positively correlated with plasma levels of sCD14, IFN-α, and TNF-α

As shown earlier, patients with lupus nephritis displayed higher plasma levels of sCD14, IFN-α, and TNF-α compared to patients without renal involvement. We evaluated plasma parameters (sCD14, IFN-α, and TNF-α) with levels of proteinuria. As shown in Supplementary Figure-1, plasma levels of sCD14 (A), IFN-α (B) and TNF-α (C) correlated positively with 24-hr urinary protein.

### Correlations of Plasma sCD14 with SLEDAI -2K

We evaluated the possibility of using sCD14 as a biomarker to assess disease severity. As shown in figure-3, plasma levels of sCD14 were significantly and positively correlated with SLEDAI scores (*P*=0.002, r=0.34). Complement components, C3 and C4, correlated negatively with sCD14 levels (C3: P = 0.003, r = -0.33; C4: P <0.0001, r = -0.50). There was no correlation with ant-dsDNA. Further, we grouped SLE patients based on the severity of the disease and compared the mean sCD14 levels among them. As shown in Figure-3B, we observed elevated levels of sCD14 in patients with high disease activity (SLEDAI scores ≥12) compared to those with mild to moderate activity (SLEDAI scores 5-12) (P=0.01).

**Figure-3.**
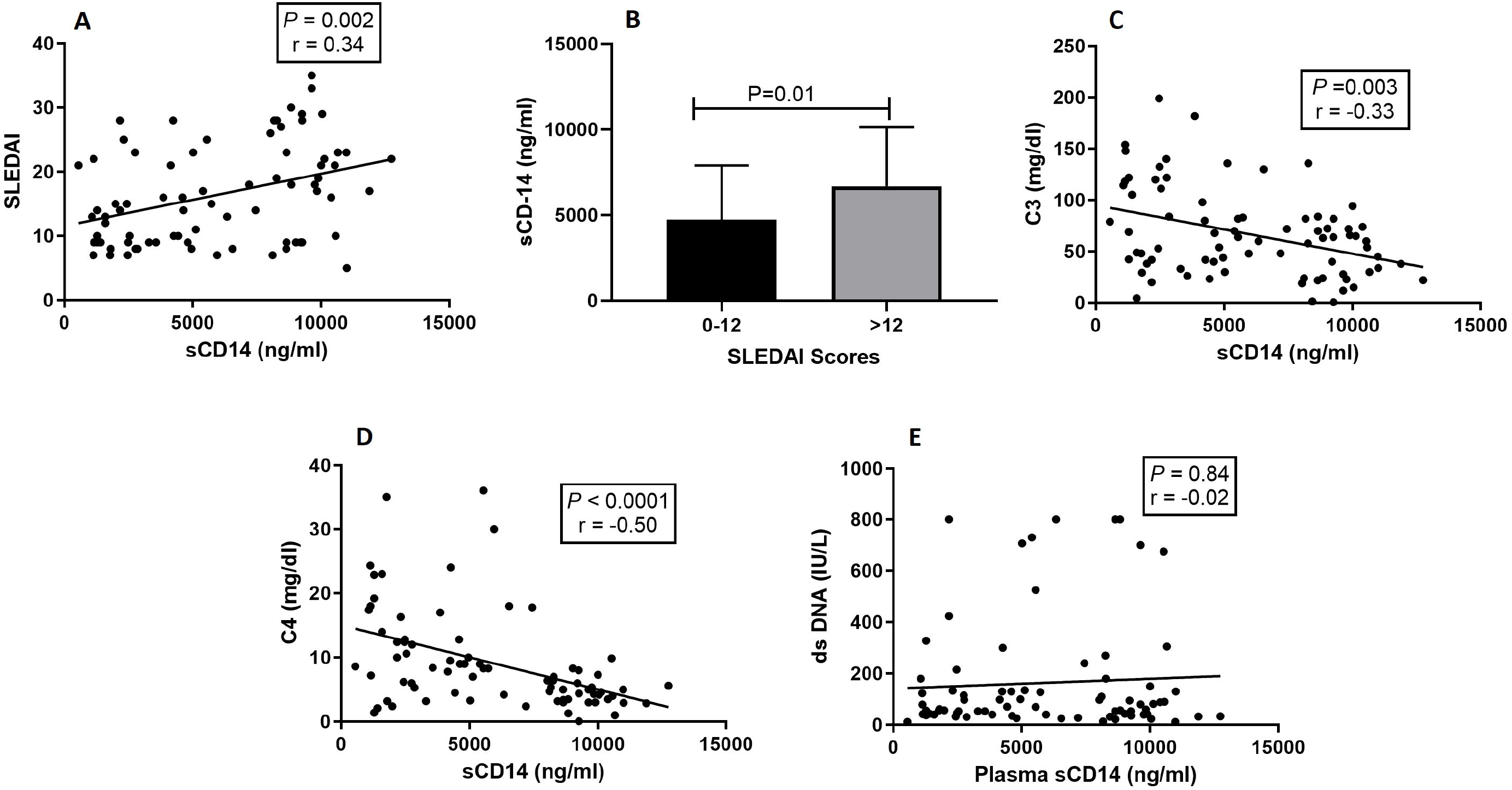
Correlation of plasma sCD14 with SLEDAI scores, C3, C4 and anti-dsDNA. Plasma sCD14 levels were quantified in SLE patients (n=78) and correlated with SLEDAI (A), C3 (C), C4 (D) and anti-dsDNA (E). A positive correlation was observed between plasma sCD14 and SLEDAI, C3 and C4. However, no correlation was noticed among sCD14 and anti-dsDNA. Further patients with severe disease phenotype displayed significantly higher sCD14 compared to moderate severity (B). Dots represent individual sample. Correlation analysis was performed by Spearman rank correlation coefficient. A *P* value less than 0.05 was considered as significant.

### Plasma levels of sCD14 correlated positively with IFN-α and TNF-α

The cytokines, IFN-α, and TNF-α have been observed to contribute to disease severity in SLE. We assessed the association of sCD14 with plasma levels of IFN-α and TNF-α, Interestingly, sCD14 levels correlated positively with IFN-α (P<0.0001, r=0.76) and TNF-α(P<0.0001, r=0.55) (Supplementary Figure-2). Furthermore, levels of IFN-α and TNF-α also showed a significant positive correlation between them (P<0.0001, r=0.51).

## Discussion

The pathogenesis of SLE is linked to the interaction between genetic susceptibility and environmental factors, notably infection, UV light, and drugs resulting in immune dysregulation. The present study demonstrates a significant association of CD14 (C-159T) polymorphism with SLE and lupus nephritis. It also provides evidence for a significant correlation between CD14 (C-159T) mutation with elevated plasma levels of sCD14, TNF-α, and IFN-α. Furthermore, we also observed that sCD14 levels can be utilized as a biomarker for assessing disease severity and lupus nephritis. The association of TNF-α and IFN-α in disease severity was reiterated. The results suggest that CD14 (C-159T) variants may be one more genetic susceptibility factors in SLE and lupus nephritis.

Association of several candidate genes has been reported in SLE over the last three decades (20, 21). However, the literature on the role or association of the CD14 promoter polymorphism in SLE is limited. In a study in Tunisian, SLE patients demonstrated an association with homozygous mutant (TT) and minor allele (T) (22). We have corroborated the above observation in our cohort: a higher prevalence of CD14 mutants (CT and TT) and minor allele (T) in SLE patients visa vis healthy controls indicating a possible contributory role in the development of SLE. We analyzed the data based on clinical phenotypes and observed a significant association with lupus nephritis. This is a novel observation. The role of CD14 mutants in the development of lupus nephritis has not been assessed. A previous study highlighted the association of T allele with higher mCD14 expression(23). And our study demonstrated the association of TT and CT genotypes with elevated sCD14 levels. Furthermore, they were also associated with elevated plasma levels of TNF-a and IFN-a; the two inflammatory cytokines that contribute to the disease pathogenesis in SLE. Interestingly, the cytokines correlated positively with plasma sCD14 levels. In summary, mutants of CD14 contribute to elevated sCD14, TNF-α, IFN-α, levels, which could be the contributing factors in disease severity and renal damage.

The number of studies is limited. Most published data indicate elevated sCD14 levels in SLE patients compared to healthy controls (8, 24, 25). Active SLE patients have been observed with higher levels of sCD14 compared to inactive disease (8, 24), and reduction of sCD14 levels have been observed post treatment. But the relationship between sCD14, IFNα, and TNFα has not been assessed.

We analyzed the data on sCD14 in lupus nephritis. The levels were significantly high compared to those without kidney involvement. Similar observations have been made in chronic kidney disease (26) and cystic kidney disease(27) but not in lupus nephritis. However, the cause and effect relationship is not known. Furthermore, higher levels of TNF-α and IFN-α were observed in lupus nephritis visa vis those without renal involvement, and they are known to be pathogenic. Elevated TNF-α and IFN-α have been observed in patients with kidney dysfunction related to various causes(28, 29). Infections are the environmental triggers that induce immune perturbation in SLE(30): CD14 recognizes a wide range of microbial pathogens that induce the production of inflammatory cytokines-TNF-α and IFN-α, and we hypothesize that this interaction, between infection and mutant CD14 results in inflammatory cytokine production that predisposes to the development of SLE and renal impairment.

The CD14 (C-159T) polymorphism is located in the promoter region of the CD14 gene. We assessed the sCD14 levels based on different genotypes of CD14(C 159T) in SLE and healthy controls. Mutants displayed higher sCD14 compared to wild type (CC), which corroborates with earlier observations (10, 11, 31). A similar observation was also noted for plasma levels of TNF-α and IFN-α. The transition polymorphism (C>T) at the promoter region decreases Sp protein binding efficiency and enhances transcription levels of CD14, leading to a higher level of sCD14 (9). Furthermore, our *in silico* analysis demonstrated the variation site as a potential area for binding of transcription factors, indicating CD14 (C-159T) mutation as a critical functional polymorphism altering soluble CD14 levels in humans.

SLE is a clinically and serologically complicated disease, and its appropriate management depends on proper diagnosis and assessment of disease severity. Several molecules and antibodies have been proposed and assessed as biomarkers for distinct clinical phenotypes, and disease activity indices like SLEDAI-2K and BILAG have been widely used to assess disease severity. There has been the inconsistency of results of biomarkers among different populations which has been attributed to ethnicity, the differential prevalence of clinical phenotypes, and the coexistence of various infectious diseases(32). Thus, a population-specific biomarker could be a useful tool for predicting disease outcome. (33). In the present study, we explored the association between sCD14 with SLEDAI-2K, C3, C4, proteinuria, and other biomarkers of disease activity such as TNF-α and IFN-α (34, 35). The sCD14 levels correlated positively and significantly with SLEDAI scores and 24 hours proteinuria; it inversely correlated with C3 and C4 levels, providing evidence that sCD14 could be a novel biomarker to assess disease activity. Interestingly, levels of sCD14 was significantly high in severe disease as assessed by SLEDAI-2K scores. This was reported in an earlier study which also observed a decrease in sCD14 levels post treatment (24).

## Conclusions

This study demonstrates a significant association between CD14 promoter polymorphism (C-159T), increased sCD14 levels with SLE, lupus nephritis, and SLEDAI-2K scores. Plasma sCD14 could be used as a novel biomarker to assess disease severity. Further studies in different populations can validate our observation.

## Data Availability

Data of the manuscript will be available upon request to the corresponding author

